# Prevention of bone loss and fractures following solid organ transplantations: Protocol for a systematic review and network meta-analysis

**DOI:** 10.1101/19013797

**Authors:** Jiawen Deng, Wenteng Hou

## Abstract

**Purpose:** Solid organ transplant (SOT) recipients can develop skeletal diseases caused by underlying conditions and the use of immunosuppressants. As a result, SOT recipients are at risk for decreased bone mineral density (BMD) and increased fracture incidences. We propose a network meta-analysis (NMA) that incorporates all available RCT data to provide the most comprehensive ranking of antiresorptive interventions according to their ability to decrease fracture incidences and increase BMD in SOT recipients.

**Methods:** We will search MEDLINE, EMBASE, Web of Science, CINAHL, CENTRAL and Chinese literature sources for RCTs, and we will include adult SOT recipients who took antiresorptive therapies starting at the time of transplant with relevant outcomes. We will perform title and full-text screening as well as data extraction in duplicate. We will report changes in BMD as weighted or standardized mean differences, and fracture incidences as risk ratios. We will use SUCRA scores to provide rankings of interventions, and we will examine the quality of evidence using risk of bias and CINeMA.

**Results:** The results of this systematic review and network meta-analysis will be published in a peer-reviewed journal.

**Conclusions:** To our knowledge, this systematic review and network meta-analysis will be the most comprehensive quantitative analysis regarding the management of bone loss and fractures in SOT recipients. Our analysis should be able to provide physicians and patients with an up-to-date recommendation for pharmacotherapies in reducing incidences of bone loss and fractures associated with SOT.

**CONFLICT OF INTEREST:** Jiawen Deng, and Wenteng Hou declare that they have no conflict of interest.

**MINI ABSTRACT:** We propose a network meta-analysis investigating the use of antiresorptive interventions to prevent bone loss and fractures in solid organ transplant (SOT) recipients. We aim to provide a comprehensive ranking of antiresorptive therapies in terms of their ability to increase bone mineral density and decrease fracture incidence in SOT recipients.

## INTRODUCTION

Solid organ transplantation (SOT) has become the standard of care for patients suffering from end-stage organ failure or organ insufficiencies in recent years. According to the United Network for Organ Sharing (UNOS), there are over 110,000 patients awaiting life-saving organ transplants in the US, with over 36,000 solid-organ transplant surgeries performed in 2018[1]. These numbers are expected to increase significantly in the near future, with worrying public health trends — such as the continuously increasing rate of cardiac failures worldwide — showing no signs of slowing down[2].

With the increasing number of transplant recipients and improvements in survival rates as a result of advancements in surgical techniques, perioperative care, and immunosuppressive therapies, improving patient’s postoperative recovery and long term survival has become ever more important. SOT patients would face many morbidity risks as a result of transplant-related consequences. The risk of osteoporosis for SOT patients, for example, has been demonstrated to be five times greater than that of the general population[3]. This may be caused by several factors, the most prevalent ones being progressive chronic disease before transplant, consequences of lifelong immunosuppression, and malnutrition[3].

There are several possible mechanisms for osteoporosis and bone loss after SOT. The majority of bone loss occur in the 6-12 months postoperative period, with a significant increase in fracture risks[4]. However, the risks of osteoporosis increase in some SOT recipient populations before the transplant. Abnormal bone formation and resorption markers are widely observed in liver transplant candidates, while altered bone metabolism is associated with hormonal abnormalities in chronic kidney disease patients[4]. In heart and lung transplant candidates, the pathophysiology and pharmacotherapies for diseases such as congestive heart failure and cystic fibrosis are also associated with an increased risk for osteoporosis pre-transplant[3, 4]. After transplant, lifelong immunosuppression is a major factor in transplant related osteoporosis. Glucocorticoids, a common immunosuppressant agent for SOT patients, activates the RANKL system which decreases bone formation and accelerate bone resorption[4]. The introduction of cyclosporine A and other calcineurin inhibitors significantly increased allograft survival and decreased organ rejection in SOT recipients; however, their effects and mechanisms on osteoporosis is still not well understood, showing inconsistent results on decreasing bone mineral density (BMD). In comparison, mammalian target of rapamycin inhibitors (mTORi) such as sirolimus and tacrolimus demonstrated an inhibition of osteoclast formation, which may promote bone growth in post-transplant patients[4, 5].

Because SOT patients are at such high risk for bone loss after transplant, osteoporosis management is very important in this population. There are several treatment options available. Calcium and vitamin D supplementation is recommended for all SOT patients even before the transplant as 91% of the patients have vitamin D insufficiencies across all end stage organ failures[6]. Beside its graft-protective effects, vitamin D improves intestinal calcium absorption, prevent hyperparathyroidism and promote osteoblast formation[3]. In addition, the effectiveness of calcitonin as an adjuvant to calcium and vitamin D has been demonstrated in renal and cardiac transplants[3]. Bisphosphonates have also been proven as robust antiresorptive agents. They maintain BMD in post-transplant patients by mediating osteoclast-related bone resorption[4] and some long term studies have demonstrated that they decrease fracture risks in SOT patients[3].

Despite the vast amount of clinical trial evidence available which supports the use of different antiresorptive drugs in transplant patients, no systematic review to date has incorporated all available randomized controlled trial (RCT) data to determine the optimal antiresorptive intervention for SOT recipients. A previous meta-analysis compared the efficacy of bisphosphonates and vitamin D analogs in increasing BMD and decreasing fracture incidences in SOT patients, however they were limited by the pairwise meta-analysis design and thus was not able to investigate the efficacy of different bisphosphonates or vitamin D analogs. In addition, they were not able to incorporate RCT data on other antiresorptive interventions, such as calcitonin[7].

Network meta-analyses (NMAs) allow the comparison and ranking of all studied interventions at once as opposed to regular meta-analyses[8]. Therefore, only a NMA can utilize all available RCT data for a given patient population. We propose to conduct a systematic review and NMA of RCTs to investigate the following research questions: What are the comparative effects (in terms of fracture incidences and changes in BMD from baseline) of different antiresorptive interventions on adult SOT recipients?

## METHODS AND ANALYSIS

We will conduct this systematic review and NMA in accordance to the Preferred Reporting Items for Systematic Reviews and Meta-Analyses (PRISMA) incorporating NMA of health care interventions[9]. This study is prospectively registered on The International Prospective Register of Systematic Reviews (PROSPERO) — *CRD42019138807*. Any significant amendments to this protocol will be reported and published with the results of the review.

### Eligibility Criteria

#### Types of Participants

We will include adult (18 years or older) SOT recipients who had received antiresorptive interventions starting at the time of transplantation. SOT is defined according to The Washington Manual of Medical Therapeutics, 34^th^ ed., as transplantation of the kidney, liver, pancreas, heart, and lung[10].

#### Types of Studies

We will include parallel-groups RCTs. If a RCT uses a crossover design, latest data from before the first crossover will be used.

#### Types of Interventions

We will include any antiresorptive pharmacotherapies used to manage bone loss and fractures. This may include (but not limited to) bisphosphonates (e.g. alendronate, risedronate, zoledronic acid), calcitonin, calcium, vitamin D or D analogs (e.g. calcitriol or alfacalcidol). If data permits, placebo and untreated (i.e. no antiresorptive treatment) will also be included as treatment arms. We will include combinations of multiple antiresorptive therapies.

### Primary Outcomes

#### Fracture Incidence

We will evaluate fracture incidences based on data collected at the latest follow-up. If data permits, we will conduct separate analyses for vertebral and nonvertebral fractures. Definitions of fractures will be defined as per individual study criteria.

#### Change in BMD

We will evaluate change in BMD from baseline, in both percentage and absolute change. BMD change must be calculated based on BMD data collected at the latest follow-up.

We will analyze BMD readings taken at the lumbar spine and femoral neck. Absolute and percentage changes in T-score and Z-score will not be included in this analysis.

### Search Methods for Identification of Studies

#### Electronic Database Search

We will conduct a librarian-assisted search of MEDLINE, EMBASE, Web of Science, CINAHL, and CENTRAL from inception to January 2020. We will use Medical Subject Headings (MeSH) terms to ensure broad and appropriate inclusions of titles and abstracts. See **Supplementary Material S1** for the sample MEDLINE search strategy.

Major Chinese databases, including Wanfang Data, Wanfang Med Online, CNKI, and CQVIP will also be searched using a custom Chinese search strategy. See **Supplementary Material S2** for the sample CNKI search strategy.

#### Other Data Sources

We will hand search the reference list of previous systematic reviews for relevant articles. We will also review clinicaltrials.gov for registered published or unpublished studies.

### Data Collection and Analysis

#### Study Selection

We will perform title and abstract screening independently and in duplicate using Rayyan QCRI (https://rayyan.qcri.org/). Studies will only be selected for full-text screening if both reviewers deem the study relevant. Full-text screening will also be conducted in duplicate. We will resolve any conflicts via discussion and consensus or by recruiting a third author for arbitration.

#### Data Collection

We will carry out data collection independently and in duplicate using data extraction sheets developed *a priori*. We will resolve discrepancies by recruiting a third author to review the data.

#### Risk of Bias

We will assess risk of bias (RoB) independently and in duplicate using The Cochrane Collaboration’s tool for assessing risk of bias in randomized trials[11]. Two reviewers will assess biases within each article in seven domains: random sequence generation, allocation concealment, blinding of participants and personnel, blinding of outcome assessment, incomplete outcome data, selective reporting, and other sources of bias. We have prospectively defined the RoB domains in **Table 1**. The overall risk of bias will be the average risk across all domains.

(Table 1: Definitions of Risk of Bias Domains)

### Data Items

#### Bibliometric Data

Authors, year of publication, trial registration number, digital object identifier (DOI), publication journal, funding sources and conflict of interest.

#### Methodology

\# of participating centers, study setting, blinding methods, phase of study, enrollment duration, randomization and allocation methods, technique for BMD measurement, technique for fracture detection.

#### Baseline Data

\# randomized, # analyzed, # lost to follow-up, mean age, sex, # postmenopausal, fracture (vertebral and nonvertebral) prevalence at baseline, baseline BMD measurements.

#### Outcomes

Final BMD measurements or percentage/absolute change in BMD from baseline, # vertebral fracture incidences at latest follow-up, # non-vertebral fracture incidences at latest follow-up.

#### Other Data

Adverse events, description of anti-diabetic and antiresorptive therapy (i.e. dosage, duration), mean follow-up.

### Statistical Analysis

#### Network Meta-Analysis

We will conduct all statistical analyses using R 3.5.1 (R Foundation for Statistical Computing, Vienna, Austria). We will perform Bayesian NMAs using the gemtc library[8]. Because we expect heterogeneity among studies due to differences in patient characteristics and study methodologies, we will use a random effects model[8]. We will use patients receiving no active antiresorptive interventions, including patients using placebo, as a reference for comparison.

For changes in BMD, we will report the results of the analysis as mean differences (MDs) with 95% credible intervals (CrIs) if all included studies utilized the same scale (e.g. if BMD changes are all reported as percentage changes). Otherwise, we will report these outcomes as standardized mean differences (SMDs) to include all available RCT data. We will calculate SMD by dividing the mean differences between treatment groups by the weighted pooled standard deviation (SD) using Hedges’ method[12]. Because SMDs are difficult to interpret for most clinicians, we will supplement our SMD results with MDs as well, considering only percentage changes in BMD. Fracture incidences will be reported as risk ratios with corresponding 95% CrIs. For studies with no fracture events, we will assign a continuity correction of 0.5[13], and we will perform sensitivity analyses to examine the effect of continuity correction factors on treatment rankings (see **Supplementary Material S3**). We will run all network models for a minimum of 100,000 iterations to ensure convergence.

If there are outcomes for which we did not gather enough information to perform a NMA, we will provide a qualitative description of the available data and study outcomes.

#### Treatment Ranking

We will use the surface under the cumulative ranking curve (SUCRA) scores to provide an estimate as to the ranking of treatments. SUCRA scores range from 0 to 1, with higher SUCRA scores indicating more efficacious treatment arms[14].

#### Missing Data

We will attempt to contact the authors of the original studies to obtain missing or unpublished data. Missing standard deviation values will be imputed using methods described in the Cochrane Handbook for Systematic Reviews of Interventions[15] using correlation coefficients (see **Supplementary Material S4**). We will perform a sensitivity analysis with different correlation coefficients to examine the effects of the coefficients on treatment rankings (see **Supplementary Material S3**), and we will comment on the consistency of coefficients across different treatments.

#### Heterogeneity Assessment

We will assess statistical heterogeneity within each outcome network using I^2^ statistics and the Cochran’s Q test[16]. We will consider an I^2^ index ≥ 50% as an indication for serious heterogeneity, and I^2^ index > 75% as an indication for very serious heterogeneity. We will explore potential sources of heterogeneity using meta-regression analyses and subgroup analyses (see **Supplementary Material S5**).

#### Inconsistency

We will assess inconsistency using the node-splitting method[17]. We will explore any indications of significant inconsistency using meta-regression analyses and subgroup analyses (see **Supplementary Material S5**).

#### Publication Bias

To assess small-study effects within the networks, we will use a comparison-adjusted funnel plot[18]. Treatment arms need to be ordered in a meaningful fashion prior to plotting the funnel plot according to assumptions of how small studies differ from larger studies. We will sort our treatment arms according to their SUCRA values with the assumption that small trials tend to favour more efficacious interventions. We will use Egger’s regression test to check for asymmetry within the funnel plot to identify possible publication bias[19].

#### Quality of Evidence

We will use the Confidence in Network Meta-Analysis (CINeMA) web application (Institute of Social and Preventive Medicine, University of Bern, Bern, Switzerland) to evaluate confidence in the findings from our NMA. CINeMA adheres to the GRADE approach for evaluating the quality of evidence by assessing network quality based on six criteria: within-study bias, across-study bias, indirectness, imprecision, heterogeneity and incoherence[20].

We will report the results of our GRADE analysis using a summary of findings table.

## DISCUSSION

The previous meta-analysis regarding the use of antiresorptive medications in SOT recipients were limited by their pairwise designs. As a result, the latest analysis did not include all available RCT data[7]. Our study aims to significantly expand upon the latest meta-analysis by incorporating the entirety of global RCT evidence available. To our knowledge, our proposed study will be the most comprehensive review to evaluate the relative effects of multiple antiresorptive agents among SOT patients using a NMA approach with multi-language search strategies.

Our review will have several strengths. First, we will extend our database search to Chinese databases for our analysis. Because of China’s immense patient population and regulations that promote pharmaceutical research, the inclusion of Chinese RCTs will help strengthen the power and precision of our analyses. Furthermore, we will use NMA techniques to analyze RCTs concerning antiresorptive pharmacotherapies. This strategy will allow us to include different bisphosphonates and vitamin D analogs as separate treatment arms, while including other interventions such as calcitonin. Lastly, we will only include RCT data, and we will use tools such as The Cochrane Collaboration’s tool for assessing risk of bias in randomized trials, CINeMA, and comparison-adjusted funnel plots to evaluate the quality of our included studies and networks. We will further use meta-regression and subgroup analyses to explore any observed heterogeneity and inconsistency, as well as using sensitivity analyses to examine the robustness of our methodology.

One major limitation of our proposed study is that Chinese clinicians may adopt different practices as Western clinicians (e.g. higher drug dosages); as a result, outcomes from Chinese RCTs may not be applicable to the Western healthcare system.

Despite this limitation, our NMA will likely be the largest quantitative synthesis assessing antiresorptive therapies among SOT recipients to date. Our study should help physicians with selecting antiresorptive regimens that are the most beneficial for the bone health of SOT recipients. Our study may also highlight promising treatment strategies that were not discussed in previous analyses, providing physicians and researchers with future research directions.

## Data Availability

All available data are presented in the manuscript and supplementary materials.

## ACKNOWLEDGEMENTS

We would like to offer our special thanks to Emma Huang, Faculty of Health Sciences, McMaster University for dedicating her time to thoroughly review our protocol.

## AUTHOR STATEMENT

JD made significant contributions to conception and design of the protocol, drafted the protocol, and substantially reviewed it. WD made contributions to the conception of the protocol, substantially reviewed it, and made revisions to the final protocol.

## REFERENCES

1. Transplant trends - UNOS. In: UNOS. https://unos.org/data/transplant-trends/. Accessed 11 Nov 2019

2. Savarese G, Lund LH (2017) Global Public Health Burden of Heart Failure. Card Fail Rev 3:7–11. https://doi.org/10.15420/cfr.2016:25:2

3. Early C, Stuckey L, Tischer S (2016) Osteoporosis in the adult solid organ transplant population: underlying mechanisms and available treatment options. Osteoporosis International 27:1425–1440

4. Lan G-B, Xie X-B, Peng L-K, et al (2015) Current Status of Research on Osteoporosis after Solid Organ Transplantation: Pathogenesis and Management. Biomed Res Int 2015:413169. https://doi.org/10.1155/2015/413169

5. Westenfeld R, Schlieper G, Wöltje M, et al (2011) Impact of sirolimus, tacrolimus and mycophenolate mofetil on osteoclastogenesis--implications for post-transplantation bone disease. Nephrol Dial Transplant 26:4115–4123. https://doi.org/10.1093/ndt/gfr214

6. Stein EM, Shane E (2011) Vitamin D in organ transplantation. Osteoporos Int 22:2107–2118. https://doi.org/10.1007/s00198-010-1523-8

7. Stein EM, Ortiz D, Jin Z, et al (2011) Prevention of fractures after solid organ transplantation: a meta-analysis. J Clin Endocrinol Metab 96:3457–3465. https://doi.org/10.1210/jc.2011-1448

8. van Valkenhoef G, Lu G, de Brock B, et al (2012) Automating network meta-analysis. Res Synth Methods 3:285–299. https://doi.org/10.1002/jrsm.1054

9. Hutton B, Salanti G, Caldwell DM, et al (2015) The PRISMA extension statement for reporting of systematic reviews incorporating network meta-analyses of health care interventions: checklist and explanations. Ann Intern Med 162:777–784. https://doi.org/10.7326/M14-2385

10. Brassil EB (2014) The Washington Manual of Medical Therapeutics, 34th ed., by Hemant Godara and Department of Medicine, Washington University School of Medicine. Medical Reference Services Quarterly 33:236–238

11. Higgins JPT, Altman DG, Gøtzsche PC, et al (2011) The Cochrane Collaboration’s tool for assessing risk of bias in randomised trials. BMJ 343:d5928. https://doi.org/10.1136/bmj.d5928

12. Hedges LV (1981) Distribution Theory for Glass’s Estimator of Effect size and Related Estimators. J Educ Behav Stat 6:107–128. https://doi.org/10.3102/10769986006002107

13. Friedrich JO, Adhikari NKJ, Beyene J (2007) Inclusion of zero total event trials in meta-analyses maintains analytic consistency and incorporates all available data. BMC Med Res Methodol 7:5. https://doi.org/10.1186/1471-2288-7-5

14. Mbuagbaw L, Rochwerg B, Jaeschke R, et al (2017) Approaches to interpreting and choosing the best treatments in network meta-analyses. Syst Rev 6:79. https://doi.org/10.1186/s13643-017-0473-z

15. Higgins JPT, Green S (2008) Cochrane Handbook for Systematic Reviews of Interventions.

16. Wiley Higgins JPT, Thompson SG, Deeks JJ, Altman DG (2003) Measuring inconsistency in meta-analyses. BMJ 327:557–560. https://doi.org/10.1136/bmj.327.7414.557

17. van Valkenhoef G, Dias S, Ades AE, Welton NJ (2016) Automated generation of node-splitting models for assessment of inconsistency in network meta-analysis. Res Synth Methods 7:80–93. https://doi.org/10.1002/jrsm.1167

18. Chaimani A, Higgins JPT, Mavridis D, et al (2013) Graphical tools for network meta-analysis in STATA. PLoS One 8:e76654. https://doi.org/10.1371/journal.pone.0076654

19. Peters JL, Sutton AJ, Jones DR, et al (2006) Comparison of two methods to detect publication bias in meta-analysis. JAMA 295:676–680. https://doi.org/10.1001/jama.295.6.676

20. Nikolakopoulou A, Higgins JPT, Papakonstantinou T, et al Assessing Confidence in the Results of Network Meta-Analysis (Cinema). bioRxiv. https://doi.org/10.1101/597047

